# Access and quality of sexual and reproductive health (SRH) services in Britain during the early stages of the COVID-19 pandemic: a qualitative interview study of patient experiences

**DOI:** 10.1101/2021.10.22.21264941

**Authors:** Raquel Boso Perez, David Reid, Karen Julia Maxwell, Jo Gibbs, Emily Dema, Chris Bonell, Catherine Heather Mercer, Pam Sonnenberg, Nigel Field, Kirstin Rebecca Mitchell

**Author notes:** **Corresponding author:** Ms Raquel Boso Perez, MRC/CSO Social and Public Health Sciences Unit, University of Glasgow, 99 Berkeley Street, Glasgow G3 7HR. joint first authors (Raquel Boso Perez, David Reid).

## Abstract

**Objectives:** Access to quality sexual and reproductive health (SRH) services remains imperative, even during a pandemic. Our objective was to understand experiences of delayed or unsuccessful access to SRH services in Britain during the early stages of COVID-19 pandemic.

**Design:** Semi-structured qualitative follow-up interviews were conducted in October-November 2020 (six months after the first UK lockdown) with participants of Natsal-COVID, a quasi-representative web-panel survey of sexual health and behaviour during COVID-19 (n=6654). Inductive thematic analysis was used to identify lessons for future SRH service access and quality.

**Setting:** Telephone interviews with participants from the general population.

**Participants:** 14 women and 6 men (24-47-years-old) reporting unmet need for SRH services and agreeing to recontact (n=311) were selected for interview using socio-demographic quotas.

**Results:** Participant experiences spanned ten different SRH services, including contraception and antenatal/maternity services. At interview, ten participants still experienced unmet need. Participants reported hesitancy and self-censorship of need. Accessing services required tenacity. Challenges included navigating inconsistent information and changing procedures; perceptions of gatekeepers as obstructing access; and inflexible appointment systems. Concerns about reconfigured services included reduced privacy; decreased quality of interactions with professionals; reduced informal support due to lone attendance; and fewer routine physical checks. However, participants also described examples of more streamlined services and staff efforts to compensate for disruptions. Many viewed the blending of telemedicine with in-person care as a positive development.

**Conclusion:** COVID-19 impacted access and quality of SRH services. The accounts of those who struggled to access services revealed self-censorship of need, difficulty navigating shifting service configurations, and perceived reduction in quality due to a socially-distanced service model. Telemedicine offers potential for greater efficiency if blended intelligently with in-person care. We offer some initial data-based recommendations for promoting equitable access and quality in restoration and future adaption of SRH services.

**SUMMARY BOX:** *What is already known on this topic:* Access to quality sexual and reproductive health (SRH) services remains imperative, even during a pandemic. In response to the threat of COVID-19, SRH services limited in-person provision, introduced social distancing and mask wearing, and expanded remote consultations and postal services. There are no published qualitative community studies in Britain exploring service-user experiences of the rapid adaption and scaling-down of SRH services in response to COVID-19.

*What this study adds:* This study provides important insights into how rapid contraction and adaptation of sexual and reproductive health services was experienced by service users. It adds the patient perspective to formal and informal learning and sharing of knowledge been practitioners and policy makers. The study highlights that difficulty accessing services, decreased quality of SRH interactions, reduced opportunity to receive informal support, and fewer routine physical checks were difficult for patients. Our data-driven recommendations – including cautious adoption of telemedicine and improving collaboration across services – have relevance across SRH services and may be useful to other primary and secondary care providers.

## INTRODUCTION

Responding to rising COVID-19 cases, the UK announced a national lockdown in March 2020, imposing stay-at-home orders and discouraging non-essential contact, including between healthcare workers and patients. To minimise SARS-CoV-2 transmission, providers rapidly adjusted how they delivered sexual and reproductive health (SRH) services through new protocols, staff redeployment, and site closures.^1,2^ Early abortion care via telemedicine was an example of successful adaptability, reducing waiting times and barriers to access^3^ and increasing service satisfaction.^4^ However, other essential SRH services such as long-acting reversible contraception (LARC) provision or asymptomatic STI screening were halted or reduced.^1,5^ Providers tried to prioritise those with most need who could not be managed remotely for in-person care.^6^

Access to SRH services remains imperative, even during a pandemic.^7^ People have a right to sexual expression, reproductive autonomy, safe childbirth, and a life free from infection.^8^ The population need for services such as routine and emergency contraception, STI testing and treatment, sexual problems advice, or antenatal care is continual. Disruptions to these services can have significant repercussions, including unplanned pregnancy, undiagnosed STIs, and sexual dysfunction.

The Natsal-COVID study sought to understand the initial impact of service reduction and reconfiguration in Britain,^9^ where SRH services are delivered by a range of providers, including general practitioners (GPs), specialist integrated sexual health services, screening programmes (e.g., chlamydia and cervical cancer) and other SRH services. Natsal-COVID Wave 1, undertaken in July-August 2020, was a quasi-representative web-panel survey designed to understand the early impact of the pandemic on SRH. It highlighted unmet need for SRH services, with 1 in 10 survey participants reporting unsuccessful attempts to access SRH services, and 1 in 5 men needing but being unable to access condoms.^10^ Other UK studies found that men who have sex with men (MSM) and young people experienced unmet need for STI testing, contraception and condom access.^11–13^ This paper describes findings from qualitative follow-up interviews with Natsal-COVID participants, to explore the experiences of unmet or delayed SRH need in the general population. Learning from patient perspectives is crucial to inform recovery and rebuilding efforts during and after COVID-19.^6^

The study aimed to explore: (1) what challenges arose for patients attempting to access and navigate SRH services during the COVID-19 pandemic, and (2) how COVID-19 protocols and reduced staffing affected patient perceptions of service quality.

## METHODS

### Study design and participants

Natsal-COVID is a mixed-method study exploring the impact of the COVID-19 pandemic on sexual behaviour, relationships, and SRH.^9^ Following the Wave 1 web-panel survey, 45 follow-up qualitative interviews were carried out to explore in-depth SRH-related issues of relevance during the pandemic. This paper draws on interviews with 20 of the 311 participants who reported unmet or delayed SRH access since lockdown in the survey and agreed to recontact. In selecting interviewees, quotas were applied to ensure variation by age, ethnicity, and region. Women were oversampled to reflect their higher use of SRH services. Ethical approval was obtained from University of Glasgow MVLS College Ethics Committee (20019174) and LSHTM Research Ethics committee (22565).

### Data collection

The research team phoned individuals who agreed to recontact, fell within the pre-specified quotas, and provided valid contact details. An introductory call explained the study and confirmed eligibility. Those interested were emailed a study information sheet and given opportunity to ask questions. Informed consent to participate was sought and recorded prior to interview. Interviews were conducted by three trained qualitative interviewers (DR, KJM, and RBP) between 02/10/2020–16/11/2020. All interviews were conducted by phone, lasting 45–90 minutes (average 70 minutes). The interview guide explored the context of help seeking, experiences of attempting to access SRH services, impact of unmet or delayed need, and attitude and experiences with telemedicine (see supplementary material). Fieldnotes (summary and reflections) were recorded after each interview. Participants were offered a £30 e-voucher for their time and contributions.

### Data analysis

Audio recordings were professionally transcribed verbatim. Transcripts were reviewed by DR, KJM and RBP for accuracy and to develop familiarity. Identifying details were removed. Data were thematically analysed,^14^ to inductively identify themes pertinent to policy and practice. Participant partner experiences were occasionally related, and these were included in analysis. Analysis was aided by NVivo 12, a CAQDAS software. Five transcripts were initially open coded by DR. These codes were reviewed by the analysis team, and a draft coding frame developed via discussion. To maximise coding consistency DR and RBP double-coded three interviews, discussing and clarifying coding uncertainties and discrepancies to develop a final coding framework that was applied to the data.

## FINDINGS

Our sample included 14 women and 6 men (Table 1). At the time of interview, ten participants reported ongoing unmet need, six had their needs partially met, and four had met their needs after some delays. Participants had commonly attempted to access more than one service. The most sought services were contraception (n=14), STI tests (n=6) and maternity/antenatal services (n=4). Our analysis identified four over-arching themes (see Table 2 for data supporting each of the themes and sub-themes described below).

**Table 1.** Sample characteristics.

|  |  |
| --- | --- |
| **Gender** |  |
| Women | 14 |
| Men | 6 |
| **Age** |  |
| 18-29 | 3 |
| 30-39 | 10 |
| 40-49 | 7 |
| **Sexual identity** |  |
| Homosexual / gay | 2 |
| Heterosexual / straight | 15 |
| Omnisexual<sup>1</sup> | 1 |
| Not reported | 2 |
| **Ethnicity** |  |
| Asian | 3 |
| Mixed | 1 |
| White | 16 |
| **Region** |  |
| England | 16 |
| Scotland | 3 |
| Wales | 1 |
| **SRH services** |  |
| Condoms and contraception | 14 |
| STI test | 6 |
| STI follow-up care | 2 |
| HIV prevention | 1 |
| HIV care | 1 |
| Sexual function and gynaecological problems | 3 |
| Fertility services | 3 |
| Advice or counselling for sexual problems | 1 |
| Maternity/antenatal services | 4 |
| Cervical screening | 3 |
| **Outcome** |  |
| Eventually met need | 4 |
| Partially met need | 6 |
| Unmet need | 10 |

<sup>1</sup> The sexual attraction to all genders.

**Table 2.** Participant Quotes Supporting Each Theme.

| **Theme** | **Example excerpt** |
| --- | --- |
| **1. Hesitation and self-censorship** | *"I didn't really want to go to the GP and bother them when other people might need to see somebody more than I do" (F18-29).* |
|  | *"Being confident enough to go and get [condoms and STI tests] in these times again, and what judgement I might receive [is a barrier] [...] I panicked that 'what if they contact a [law enforcement] service and I get fined'." (M18-29)* |
| **2.1 Navigating inconsistent information and changing procedures** | *"They kept messing her about saying to her, 'come', then 'don't come', then 'come', then 'don't come' [to have a weight-check in order to access contraception] (M30-39).* |
|  | *"We were basically the tennis ball between these two departments [the GP and midwifery]" (M30-39)* |
|  | *"I was left helpless, stuck with menopause symptoms" (F40-49).* |
| **2.2 Navigating past gatekeepers** | *"It just made me feel like I didn't matter" (F40-49).* |
|  | *"I can't get a referral [...] I was honestly going around in circles all the time. In the end because I was getting turned away all the time by the...on phone calls, I'd just think well what's the point, there's no point in me even ringing up. In the end I just suffered on really." (F30-39)* |
|  | *"There's a special ward for mums that are having problems. So, it's like an emergency type of thing, so that was quite difficult to access [...] we've got family who are doctors, so they know how these work [...] So, we were able to understand what the system was like, but I think people who don't have people in the know it must be a lot more difficult." (M30-39)* |
| **2.3 Navigating inflexible appointment systems** | *"Because I was driving, I couldn't take the phone call and I think then I had to phone up the next day and I had to go through the same process, I had to say that if I don't pick up the phone I'm driving, please phone back". (F40-49)* |
| **3.1 Reduced privacy** | *"You have to talk through an intercom, from the outside [of the GP]. And everybody now knows what's wrong with you, [...] it's on the street, as well, so anybody walking past can just hear what you're saying." (F40-49)* |
| **3.2 Decreased quality of interactions with professionals** | *"The person who did my stuff had a mask, but we didn't talk as much as we would have. They just got down to the questions [...] Normally when I go to these places we have like a little discussion, like a little pep chat". (M18-29)* |
|  | *"There's always a bit of a delay on the video, and because it's a bit stilted, and because I'm hard of hearing, I can't always lip read." (F40-49)* |
| **3.3 Reduced informal support due to lone attendance** | *"As good as all the staff are, if you're getting bad [pregnancy] news in a situation like that it is going to cause you a lot more stress if you don't have someone with you." (F30-39)* |
|  | *"He feels less involved. He's not there to see and hear for himself sort of what's going on. He's having to sit and wait for me to relay everything." (F30-39).* |
| **3.4 Fewer 'best practice' routine physical checks** | *"I'm totally frustrated because everybody should have the same right to be checked [...] they should have some period for people who just want to get [STI] tested." (F18-29)* |
| **3.5 Improved efficiency** | *"There were a few silver lining moments. We didn't have to wait that long to go into our appointment, the appointments themselves were a lot faster. We still got the good service from the midwives". (F30-39)* |
|  | *"They did all the tests there and then, but they had to send them off and then they phoned me, I think, a week later to say that everything was fine apart from the chlamydia which was positive and that they would...they posted the antibiotics out, so that was quite easy actually, it arrived in the post." (F40-49)* |
| **4. Attitudes toward the continuation of telemedicine** | *"The phone probably is easier because you're not having to look at somebody and you don't get the nervousness [...] if you're suddenly kind of face-to-face with somebody that looks quite official, that might make you clam up." (M30-39)* |

### 1. Hesitation and self-censorship

Participants frequently discussed their hesitation to use services. Some feared contracting COVID-19 through clinic attendance. Many downplayed their needs relative to others, particularly people with COVID-19, and did not wish to burden an already pressured health system. Several participants self-censored their needs after encountering barriers to access like long queues or difficulty booking appointments, others due to fear of service providers disapproval for being sexually active when contact between households was restricted.

### 2. Navigating new and shifting service configurations

Participants typically found services harder to access than before. Several found that SRH services were closed, on pause, or not accepting referrals.

#### 2.1 Navigating inconsistent information and changing procedures

While making enquires, being triaged, or attending appointments, eleven participants described dealing with inconsistencies in information or sudden changes to services. One detailed how his partner was told she required a weight-check to access contraception, yet her surgery was not offering in-person visits for weight-checks. Another described phoning over ten times to access antenatal care, as their GP had advised that they could self-refer to midwifery, whilst midwifery insisted that the GP needed to refer them. Several described how in-person appointments were cancelled or carried out remotely, leaving them uncertain of when they could access treatment.

Changes to services reduced participants’ sense of choice and autonomy, resulting in some accepting less preferred alternatives. Service changes were perceived by many as insurmountable—particularly for those seeking face-to-face appointments, LARC, asymptomatic STI testing, or fertility treatments. Those seeking contraception for reasons other than preventing pregnancy (e.g., managing menopausal symptoms) felt their needs were deprioritised.

#### 2.2 Navigating past gatekeepers

Participants highlighted new layers of triage (added stages before consultation) and increased gatekeeping. Reaching their desired healthcare professional took longer or proved harder. Gatekeepers included receptionists, nurses, doctors, and to a lesser degree automated online/telephone systems whose remits covered assessing need and giving information on changes to service configuration. These gatekeepers arranged appointments, denied access to, or referred onward depending on availability and the service sought. In-person care was notably difficult for some to access, with one participant describing it as “*impossible*”. Several participants were told the service they needed was “*on hold*” or were referred elsewhere. Initial access was often hardest; those requiring multiple healthcare interactions (e.g., STI treatment or antenatal services) experienced less difficulty once they were in the system. Those with more healthcare knowledge and financial resources discussed how these minimised barriers to access, one participant discussed booking “*a private scan that allowed partners to come in*” (M30-39).

Various participants described the negative impact of service providers not following-up after a failed access attempt, putting the onus back on the patient. Even with persistence and tenacity, not all were able to overcome barriers, including one woman suffering intense gynaecological pain.

#### 2.3 Navigating inflexible appointment systems

Typically, walk-in or same day services were replaced with triage and appointment systems. One participant described queues and being turned away when she arrived for her pre-booked appointment. Several participants were frustrated by the increased need for forward planning. A less flexible service made it difficult for some participants balancing other commitments (e.g., childcare) to access healthcare. Participants navigating phone appointments discussed the difficulty of not knowing when a clinician would contact them and the challenge of needing to restart the process if they missed their call-back.

### 3. Experiencing a ‘socially-distanced’ and reduced service

Twelve participants (or their partners) eventually attended the service they required. They described their experiences of engaging with services operating under COVID-19 protocols and reduced staffing.

#### 3.1 Reduced privacy

COVID-19 safety protocols impacted participants’ privacy. Some could not openly discuss their SRH needs within their home during remote consultations, whilst others discussed being expected to disclose sensitive information in settings that did not feel private (e.g., queuing outside the GP alongside other patients).

#### 3.2 Decreased quality of interactions with professionals

Several participants reported that changes to the tone, pace, and nature of clinical interactions resulted in less opportunities to address problems, ask questions, or have spontaneous discussions. Some felt communication quality decreased during remote appointments. One participant discussed how his blood-test result for HIV management “*wasn’t very well explained*” (M30-39) compared with previous in-person consultations; another described rushed antenatal appointments as “*five minutes on the phone and then you’re done for a month*” (F30-39). Some felt staff had less welcoming attitudes, aligning with the more formal patient interaction protocols.

Some participants were disappointed they could not speak to a familiar and trusted healthcare worker. Others missed the reassurance of in-person appointments. The substitution of in-person with remote consultation was experienced as less supportive for those more concerned for their health. Remote consultations were also difficult for those with accessibility needs, such as those deaf or hard-of-hearing.

#### 3.3 Reduced informal support due to lone attendance

For some participants, challenges were compounded by having to attend appointments alone, without anyone to support, ask questions, or help remember information on their behalf. This concern was most prominent for those navigating fertility or antenatal/maternity services.

Pregnant couples discussed how the distress of lone attendance largely impacted women; one partner described his pregnant wife as “*traumatised*” from dealing with pregnancy complications on her own. Participants perceived antenatal care as a shared SRH need. Lone attendance rules left men feeling disengaged, uncertain what of was happening, and less equipped to emotionally support their partner.

#### 3.4 Fewer ‘best practice’ routine physical checks

Reduced in-person contact was perceived to reduce best practice checks such as blood-tests when starting medication or weight-checks for contraception. Many participants described how they were unable to continue SRH practices they saw as routine or positive, such as asymptomatic STI screening or cervical smear testing.

#### 3.5 Improved efficiency

A few participants noticed staff making extra effort to be friendly and reassuring. Furthermore, some changes implemented were experienced positively (e.g., shorter appointments and waiting times, less crowded waiting areas). Several participants discussed the benefits of a blended approach that combined telemedicine’s convenience with in-person care. One participant spoke about the benefits of liaising on the phone and postal delivery of antibiotic treatment for her STI infection.

### 4. Attitudes toward the continuation of telemedicine

Many viewed telemedicine—if used to complement in-person services—as potentially adding quality. Participants felt it could bring convenience to triaging, referrals, information giving, and postal medication delivery. Telemedicine could allow people to save time and money normally spent on childcare or travel. Many appreciated being able to discuss sensitive health needs at home, a familiar and non-clinical environment. Telephone (but not video) appointments particularly lessened perceived concerns around stigma of accessing SRH services. Those critical of telemedicine worried it would duplicate consultations or result in burdensome arrangements where calls were not returned, and phone line staffing was variable. Several participants worried that explaining symptoms, physical examination, and testing may be impossible remotely.

## DISCUSSION

These findings complement previous research on how COVID-19 has disrupted and impacted SRH,^10,11,15^ highlighting how unmet or delayed need might have resulted in worsened SRH outcomes, undermined reproductive choice, weakened preventative practices, and increased distress over SRH. Participants faced barriers due to changing regulations around healthcare access, inconsistencies between and within services, the pausing of various services, and difficulty navigating remote healthcare. Consequently, participants required health literacy and tenacity to access SRH services, which raises concerns about the amplification of pre-existing health inequalities given the social patterning of health literacy,^16,17,18^ and existing barriers to SRH access.^19–22^

Telemedicine represents perhaps the most significant adaptation deployed by providers to mitigate the pandemic’s effects on healthcare. This study provides detailed qualitative evidence of the mixed picture around its acceptability and suitability. Corroborating wider research on adults’ experiences of SRH telemedicine,^3,23–25^ some Natsal-COVID participants held positive attitudes toward telemedicine, especially when complementing in-person care. This differs from findings amongst young people who hesitate to access remote SRH care.^11,26,27 28^However, our participants were less satisfied with telemedicine during more sensitive and emotional consultations, highlighting the limits of remote provision.^28^ Considering privacy and anonymity of SRH patients is crucial.^19,26,29^ Some participants worried that telemedicine might alert others in their household or community to their SRH needs. However, research has also highlighted the potential for telemedicine to promote privacy and anonymity in certain contexts, such as abortion care.^23^ Given their varying needs and preferences, our data provides evidence that SRH patients should have the option of in-person, over-the-phone, or video appointments to meet varying needs and preferences. This will require investment in training and equipment to ensure high-quality remote services.^30^

### Strength and limitations

We purposively interviewed participants who had tried but failed to access services. Quota-sampling from a quasi-representative population survey permitted us to include experiences from participants who varied by age, gender, ethnicity, and region. Although our enquiry followed a holistic framework of SRH, we were limited to issues experienced by our participants; services not covered included abortion and sexual assault services. Our study was unable to explore in depth the impacts of *delayed* access to specific services. Given the time between reporting of help-seeking and interview, we could not exclude potential for recall bias. This study aimed to highlight challenges, and participants were recruited accordingly. Thus, results likely underrepresent positive experiences, such as professional staff’s enormous efforts during this challenging time. Given participants’ digital recruitment, we may not have captured experiences of those without access to remote services, for example, due to language barriers, learning difficulties, or socio-economic factors. Finally, as with all qualitative research, our study draws on a small sample to capture a range of experiences of SRH access; it is not intended to be generalized or quantified.

## CONCLUSION AND RECOMMENDATIONS

Demand for services may increase due to a backlog of delayed help-seeking and the possibility of increased compensatory risk behaviours post-pandemic. Our study provides a general population perspective to complement service-user studies and quality improvement studies, offering recommendations for future practice derived from actual and potential patients in the community. Based on our qualitative data, and in discussion with the study team and clinical colleagues, we set out a draft set of recommendations for consideration by service providers and policymakers (Table 3). These recommendations link directly to the data and represent experience-learning from this unprecedented period to support a strong recovery, innovative and streamlined services in future, and resilience during future pandemics. However, implementing these recommendations must centre the wellbeing of NHS staff, on whom the pandemic has taken a significant toll. Long term adequate investment is crucial to safeguard staff and patient wellbeing.

**Table 3.** Recommendations.

| **Service Delivery Aspect** | **Preliminary recommendations suggested from the data** | **Link to data theme** |
| --- | --- | --- |
| **Recovery phase** | In encouraging re-engagement with services, reassure patients of the legitimacy of their needs and staff's non-judgemental attitude. This applies particularly to those reporting risk behaviour during the pandemic. In the longer term – and once initial backlogs are cleared – targeted campaigns may be required to encourage re-engagement with cervical screening or asymptomatic STI testing and training for providers. | 1. |
|  | Review gatekeeping functions established during the pandemic to ensure remaining triage systems do not create additional barriers for patient access. | 2.2 |
| **Innovations to 'usual practice'** | Cautiously adopt telemedicine where it can enhance convenience and enable prompt testing, diagnosis and treatment responsiveness. It should be considered in addition to face-to-face services. Effort will be required to avoid telemedicine's unnecessary bureaucratisation, duplication or the creation of added barriers to patient access. It will be crucial to avoid exacerbating inequalities in access and digital exclusion. | 4. |
|  | Patient preferences for phone, video, or in-person consultation should be respected where possible, as needs, preferences and concerns vary. | 4. |
|  | Ensure gatekeepers are aware of their role within a triage system, so that they can supply patients with as much accurate, up-to-date and reassuring information as possible. | 2.2 |
| **Service delivery during a pandemic** | Prioritise cross-sector collaboration to avoid confusion over triage, particularly between pharmacies, GPs, and specialist SRH providers. | 2.1 |
|  | Set up agile and accessible mechanisms for sharing learning and good practice. The COVID-19 resources and regular meetings established by BASHH (British Association of Sexual Health and HIV) are a good example of this. | ALL |
|  | Ensure that information (e.g. booking systems, opening hours) is continuously updated to avoid confusion for patients. | 2.1 |
|  | Allow patients to be accompanied during pregnancy/antenatal services, during other emotionally demanding appointments, or to help with access needs (e.g., language, disability, vulnerability) wherever possible. | 3.3 |
|  | Establish safe ways to help patients feel comfortable in clinic to compensate for measures such as masks, or socially distanced consultations. Small gestures (such as a warm greeting) may be 'quick wins' reducing stress both for the patient and professional and ensuring a more personable service. | 3.2 |
|  | Continue with remote provision where possible, practicable and acceptable, ensuring staff are appropriately trained and supported to provide it. | 4. |

## Supporting information

Interview Schedule

SRQR Checklist

## Data Availability

Natsal-COVID qualitative interview data is available upon request from researchers. Natsal-COVID quantitative survey data is soon to be available from the UK Data Archive.

## Contributors

RBP and DR contributed equally to this paper. The paper was conceived by KRM and DR, with further discussions with RBP, KJM, JG, ED, CB, CHM, PS and NF. RBP, DR, KJM and KRM wrote the first draft, with further contributions from JG, ED, CB, CHM, PS and NF. Qualitative interviews were undertaken by DR, RBP and KJM. Coding undertaken by DR and RBP. Analysis of the qualitative data was carried out by RBP, DR, KJM and KRM. PS and CHM are Principal Investigators (PIs) on Natsal and NF and KRM are PIs on Natsal-COVID. All authors contributed to data interpretation, reviewed successive drafts, and approved the final version of the manuscript.

## Acknowledgments

We want to thank the study participants and Margaret Blake and Reuben Balfour (Ipsos MORI). We thank Claudia Estcourt, Clinical Professor of Sexual Health & HIV, and Honorary Consultant in Sexual Health in NHS Greater Glasgow & Clyde’s Sandyford Services, with who we discussed the paper’s recommendations. We thank Malachi Willis, Research Associate and the MRC/CSO Social and Public Health Sciences Unit, who provided proof reading and copy editing. Finally, we acknowledge members of the National Institute for Health Research Health Protection Research Unit (NIHR HPRU) in Blood Borne and Sexually Transmitted Infections (BBSTI) Steering Committee in securing funding for this NIHR HPRU: Professor Caroline Sabin (HPRU Director), Dr John Saunders (PHE Lead), Professor Catherine Mercer, Dr Hamish Mohammed, Professor Greta Rait, Dr Ruth Simmons, Professor William Rosenberg, Dr Tamyo Mbisa, Professor Rosalind Raine, Dr Sema Mandal, Dr Rosamund Yu, Dr Samreen Ijaz, Dr Fabiana Lorencatto, Dr Rachel Hunter, Dr Kirsty Foster and Dr Mamooma Tahir.

## Patient and Public Involvement

Patients or the public were not involved in the design, or conduct, or reporting, or dissemination plans of our research.

## Ethics approval

Ethical approval was obtained from University of Glasgow MVLS College Ethics Committee (20019174) and LSHTM Research Ethics committee (22565).

## Transparency statement

This manuscript is an honest, accurate, and transparent account of the study being reported, no important aspects of the study have been omitted.

