## Supplementary material for "Access and quality of sexual and reproductive health (SRH) services in Britain during the early stages of the COVID-19 pandemic: a qualitative interview study of patient experiences": Interview Schedule

**Natsal Covid-19 study. Qualitative Interviews Topic Guide**

### GROUP TWO: Participants with unmet SRH needs

**INTRODUCTION**

- Ensure participant has read the Participant Information Sheet and consent form
- Explain the study purpose and confirm permission for digital recording.
- Reiterate re data confidentiality and storage/use of data
- Remind participant that we do not have access to their survey responses and so may end up covering some similar ground in this interview
- Offer to answer any questions (repeat this offer at the end of the interview as well).
- Remind participant that they are not obliged to answer questions and can end the interview at any time.
- Since Covid a lot of people have been spending more time at home. Talking about health and relationships can be quite personal. Do you have any concerns about family, partners or friends overhearing you? Is there somewhere in your home where you can talk privately / you feel comfortable talking about these topics?
- If the connection should fail for any reason, I will call you back. Please wait for me to call you.
- **RECORD verbal consent**
    - Have a first audio recording at the start of the interview where people state their name and that they say that they have consented over email and are happy to take part.
    - Start a second audio recording for the interview itself.

**Socio-demographic and lockdown arrangements**

- Confirm:
    - age
    - gender
    - ethnicity
    - sexual orientation
    - region (SE England, SW England, Midlands, North England, Wales, Scotland)
    - location (rural, peri-urban, urban)
    - current partnership and family status (any children; their ages and current location)
    - occupation (public facing, continued, homeworking, furloughed, unemployed).
- Ask about **household in which they were locked down**:
    - **number of others** in household; gender, age, occupation (public facing, continued, homeworking, furloughed, unemployed)
    - **relationship to others in household**;
    - details of **any moves** from one HH to another over period of full/partial lockdown

**Views on Covid-19 and social restriction measures**

- What are your thoughts on Covid-19?
    - Feelings about whether social restrictions are justified or not
    - General level of concern and nature of concerns (re virus itself, economy, wellbeing, impact on health services and education, concerns about friends/relatives)

- Perceived risk or experience of covid-19 to self and others in the household?

**Covid-19 and personal life**

Covid-19 has changed life for many people. What do you see as the most important gains and losses (or positives and negatives) for you?

*Let participants speak and use the bullet point below as probes. Ask whether changes are perceived as positive, negative, both.*

- Additional time on hands/less time
- Changes to financial situation
- Changes to living arrangements
- Changes to family life/caring responsibilities
- Changes to mental and physical health?
    - Mental wellbeing
      Changes to exercise, diet, smoking, drinking etc.
- Changes to social life (more or less interested in interacting with people you are close with (friends/family/others)? What has that meant in practice for your interaction with them? (prompt: in person/ online?)

**Sex and relationships in general**

*I'd like to ask you a bit about your sex life/sexual relationships more generally. We may have covered some of this already but I'd just like to check in. As a reminder you don't need to answer anything you feel uncomfortable*

- Has your sex life changed since March?
    - (probe: satisfaction, thoughts and feelings, partnered activity, enjoyment, online activities, new practices or partners, frequency, anxiety, risk, pressures)
- How do you feel about these changes?

**Unmet sexual health needs**

In the next section we will talk about your experiences of trying to get a range of sexual and reproductive health services. These are things that you might have considered, tried to get, accessed through different means than usual, or which you may have had problems getting since the beginning of lockdown.

I'm going to read out a list of services. Let me know if any are relevant to you and if you would be happy to discuss them.

While it is useful to our research, you don't have to talk about anything you feel uncomfortable about and we can also stop and move on to something else at any time.

- Condoms and contraception
- STI testing and care
- HIV prevention and treatment
- Fertility services/advice
- Maternity/antenatal services
- Cervical screening (smear test/pap test)
- Abortion/Pregnancy termination services
- Advice or counselling for sexual problems

- Relationship support services/advice
- Sexual assault/rape support services or helplines

Were there any other sexual or reproductive health services that I haven't mentioned that you thought about or tried to access since lockdown began? Would you be happy to talk about them? (list and not if happy to discuss)

**<u>Questions on services considered, sought, used, not used or alternatives used.</u>**

*(make sure you come back to any services above considered relevant and happy to talk about)*

You mentioned _______ were/was relevant to you since the beginning of lockdown. **Can you tell me what was going on for you during that time?**

**I'd like to talk in detail about your thought processes and the steps you took (or didn't take)**

**Probes.**

**<u>Before seeking help</u>**

- Please tell me a little about the situation that led you to want to seek a service (probe for severity/urgency of need)
- Have you sought help previously for this need? (if yes, please tell me briefly about your past experiences of getting this help – and feelings about that)
- Which particular service(s) did you consider and why? [reasons for discounting other services]
- Did you talk to anyone or get advice or look up anything online prior to seeing formal help?
- What expectations did you have about how easy or difficult it would be to access the service?
- Prior to accessing/attempting to access the service, did you have any concerns? Or thoughts about what might happen if you didn't get the service?
- What other things did you do or consider doing before or alongside seeking formal help? (informal sources of support, ignoring problem etc)
    - What happened; how did you feel about what happened?

**<u>Attempting to access formal help</u>**

- Please tell me about the process of trying to access help/service/commodity: How did you try to get [the service]?
    - Probe for the process of seeking help (e.g. getting in contact with services; finding out information; process of setting up appointment; delays – waiting times; alternative routes for getting service.
    - Did anything make it harder for you to access the service? What was this?
    - Did anything make it easier for you to try to access the service? (e.g. other sources of help)
    - What actions were you able to take yourself? Anything that you wished to do that you weren't able to?
    - If help-seeking unsuccessful, what other things did you do or consider doing? (e.g. informal help, changing behaviour, finding other ways of coping)
    - Did these difficulties in attempting to access help have consequences for you in terms of your sex life/relationship?

- **If previously sought help for this issue,** what are the key changes to accessing services that you noticed and how did you feel about them?

<u>**Experiences and feelings about getting formal help (for P who did get help)**</u>

- Please tell me about your experience of receiving help? (desired level of support received? Sense of agency/autonomy, choice/options, efficiency, convenience, quality)
- If **telemedicine** accessed: how did you feel about phone/video contact? (likes, dislikes; benefits, drawbacks) How would you feel about these services becoming standard practice in future?
- **If previously sought help for this issue,** what are the key changes to receiving/experiencing services that you noticed and how did you feel about them? (changes to: desired level of support received? Sense of agency/autonomy, choice/options, efficiency, convenience, quality)
- Were there any changes to [the service] during lockdown that worked well for you?
    - Would you keep any of them in the future?
- Were here any changes to [the service] that didn't work for you? What were they?
- Did these changes (if any) to service provision (e.g. delays) have consequences for you in terms of your sex life/relationship?
- If you could make any changes to [the service] what would these be? (probe for both during a pandemic, and in the longer-term)
    - Is there anything that could make it work better for you?
    - *If they are struggling to come up with ideas offer prompts of things they might want changed:* ways of contact the service, seeing a different staff or role (e.g. nurse of doctor), staff attitude, waiting times, location of service, in person or telemedicine, staff knowledge, costs, time.

<u>**If no service/help accessed:**</u>

- **Can you tell me how (ignoring, delaying, not getting or using an alternative) service has affected you since lockdown began?** (probe for negative and positive effects on health, sex and sex drive, well-being, relationships, economic/resources and social effects).
- Do you think this has been a common experience for other people in your situation?
- Do you have any thoughts about how the government could make SRH services more easily accessible during a pandemic? What about more generally in future?

**Before we finish, I'd just like to ask you some <u>general questions</u> (also for respondents with few services considered, used or unmet)**

- **How do you feel about sexual and reproductive health services that are provided online or through telephones?**
  I'm thinking about things such as ordering condoms or STI tests by mail for postal delivery, filling in health checks online and consultations with health professionals or counsellors by telephone or by video? It might also include text reminders as well as phone apps and devices that might send information to medical professionals

- First can I check whether you have experience of using any online or over the phone SRH services before the covid lockdown? Can you tell me about that (type of service used and feeling about experience)

- If no experience of telemedicine: What do you feel would be the drawbacks and benefits of telemedicine for your personally?

- For both: How would you feel about a general move towards telemedicine or services provided online in future? (which aspects are acceptable/desired; ideas on best/most appropriate modes of delivery)

- **It has been suggested that other venues could be used to deliver some services, for example pharmacies, maternity and abortion services. How would you feel about using these?**
    - What other services / venues would you be happy using for SRH services / advice?
    - Have there been changes to sexual health services resulting from lockdown that you have liked and would like to see continued or expanded?
        - What makes you say that?
    - Is there anything that you think might make accessing services in these ways easier/harder?

**<u>Wrap-up question</u>**

- Thinking about covid and the current situation, what do you think life will look like in a year's time?
    - (prompt) Do you think life will have gone back to "normal"?

**END INTERVIEW:**

- Thank participant;
- Ask if they would like to be informed about the outcome of the project – if so ask them for an email address to add to the participant findings mailing list;
- Reiterate re data storage and use;
- Ask if any questions
